# Clinical evaluation of a low-coverage whole-genome test for detecting homologous recombination deficiency in ovarian cancer

**DOI:** 10.1101/2023.12.07.23299362

**Authors:** Romain Boidot, Michael G.B. Blum, Marie-Pierre Wissler, Céline Gottin, Jiri Ruzicka, Sandy Chevrier, Tiffany M. Delhomme, Jérome Audoux, Adrien Jeanniard, Pierre-Alexandre Just, Philipp Harter, Sandro Pignata, Antonio González-Martin, Christian Marth, Johanna Mäenpää, Nicoletta Colombo, Ignace Vergote, Keiichi Fujiwara, Nicolas Duforet-Frebourg, Denis Bertrand, Nicolas Philippe, Isabelle Ray-Coquard, Eric Pujade-Lauraine, the PAOLA-1/ ENGOT-ov25 Study Group

## Abstract

**Background:** The PAOLA-1/ENGOT-ov25 trial showed that maintenance olaparib plus bevacizumab increases survival of advanced ovarian cancer patients with homologous recombination deficiency (HRD). However, decentralized solutions to test for HRD in clinical routine are scarce. The goal of this study was to retrospectively validate on tumor samples from the PAOLA-1 trial, a decentralized HRD test based on low coverage shallow Whole Genome Sequencing (sWGS).

**Methods:** The study comprised 368 patients from the PAOLA-1 trial. The sWGS test was compared to the Myriad MyChoice HRD test (Myriad Genetics), and results were analyzed with respect to Progression-Free Survival (PFS).

**Results:** We found a 95% concordance between the HRD status of the two tests (95% Confidence Interval (CI) 92%-97%). The Positive Percentage Agreement (PPA) of the sWGS test was 95% (95% CI; 90%-97%) like its Negative Percentage Agreement (NPA) (95% CI; 89%-98%). Only 1% (95% CI; 0-3%) of its results were inconclusive. In patients with HRD-positive tumors treated with olaparib plus bevacizumab, the PFS Hazard Ratio (HR) was 0.38 (95% CI; 0.26-0.54) with sWGS and 0.32 (95% CI; 0.22-0.45) with the Myriad assay. In patients with HRD-negative tumors, HR was 0.98 (95% CI; 0.68-1.41) and 1.05 (95% CI; 0.70-1.57) with sWGS and Myriad tests. Among patients with BRCA-wildtype tumors, those with HRD-positive tumors, benefited from olaparib plus bevacizumab maintenance, with HR of 0.48 (95% CI: 0.29-0.79) and of 0.38 (95% CI: 0.23 to 0.63) with sWGS and Myriad test.

**Conclusion:** The SeqOne sWGS assay offers a clinically validated approach to detect HRD.

## Introduction

According to the World Health Organization’s Global Cancer Observatory, the number of ovarian cancer cases is predicted to rise to 442,721 by 2040, marking a 42% incidence increase from 2020 (1). This cancer is usually diagnosed at late stages and patients with advanced ovarian cancer often have poor prognosis, with five-year survival rates in high-income countries varying from 36 to 46% (1). In France, Net survival rates of 42% were reported for patients diagnosed with ovarian cancer between 2010 and 2015 (2). However, the development of Poly-ADP Ribose Protein (PARP) inhibitors has contributed to increasing the efficacy of maintenance treatments, as shown in several phase 3 clinical trials (3–6). These inhibitors trap the PARP enzyme that is involved in the repair of single-strand DNA breaks, inhibiting the default repair pathway in cells with homologous recombination deficiency (HRD) (7,8). In the PAOLA-1/ENGOT-ov25 phase 3 trial investigating first-line maintenance treatment for advanced ovarian cancer, the maximum benefit of olaparib plus bevacizumab maintenance was observed in patients with BRCA1/2-mutant and/or homologous recombination-deficient tumors (9). HRD testing and BRCA testing are therefore recommended for an optimal management of advanced ovarian cancer patients.

In the US, the MyChoice CDx HRD test (Myriad genetics) has been approved by the FDA for HRD testing in ovarian cancer, and it is recommended by ASCO guidelines (10). In other countries, such approved companion tests do not exist, and HRD and *BRCA* testing may be performed in local diagnostic labs instead of a centralized testing facility. Therefore, it is important to provide clinically-validated in-house HRD testing in these countries in order to better guide decisions to prescribe PARP inhibitor (11). In addition, integrating HRD testing into clinical practice worldwide also requires affordable and time-efficient solutions (12).

In recent years, several tests have been developed to detect HRD. Some of them are based on the whole-genome sequencing or gene panels to capture mutational signatures and genomic scars, while others rely on functional tests assessing the ability of tumor cells to perform homologous recombination (13–17). However, clinical validation studies of decentralized commercially available HRD tests are scarce: to our knowledge only two such assays have been studied, the decentralized version of the Myriad myChoice test (11) and Sophia Genetics solution that relies on a deep learning model to predict genomic instability (18). Several laboratory developed tests (LDTs) from academic institutions have also been evaluated based on the PAOLA-1/ENGOT-ov25 trial data (19–21). However the large access to these academic-based HRD tests remains a challenge.

SeqOne Genomics has developed an HRD testing procedure that relies on shallow Whole Genome Sequencing (sWGS) to capture genomic instability with limited genomic coverage. In this study, we aimed to evaluate the clinical utility of the sWGS SeqOne HRD test in advanced high grade epithelial ovarian carcinoma patients, based on a retrospective analysis of tumor DNA and clinical data of 368 patients included in the PAOLA-1/ENGOT-ov25 trial.

## Materials and Methods

### Patients and tumor material

The study comprised 368 patients with high-grade ovarian cancer (FIGO stage III to IV) enrolled in the PAOLA-1/ENGOT-ov25 phase 3 trial (NCT02477644), for whom enough DNA tumor was available. A complete list of patient eligibility criteria is provided in the description of the PAOLA-1 trial (9). The clinical data of the patients and the DNA extracted from the tumor material were provided by the clinical and translational departments of the ARCAGY-GINECO collaborative group.

### DNA sequencing

For library preparation, 200 ng of DNA from solid tumors were enzymatically fragmented and prepared with the SureSelect XT HS2 DNA Reagent Kit (Agilent Technologies, Inc.) following the manufacturer’s instructions. Paired end sequencing was performed on a NovaSeq 6000 device (Illumina, Inc.) to determine *BRCA* status. Shallow Whole Genome Sequencing was also performed on material obtained after the first PCR amplification that preceded hybrid capture enrichment using a NovaSeq 6000 device. An additional reproducibility experiment was conducted on 94 PAOLA-1 samples which were analyzed by paired end sequencing on a NextSeq500/550 device (Illumina, Inc.). For *BRCA* testing, DNA was captured using the Agilent HRR 17 panel that targets *TP53* and seventeen genes involved in the Homologous Recombination Repair (HRR) pathway: *ARID1A, ATM, BRAF, BRCA1, BRCA2, BRIP1, CDK12, CHEK1, CHEK2, FANCA, FANCL, NBN, PALB2, PIK3CA, RAD51C, RAD51D*, and *ZNF276*.

### HRD test and *BRCA* variant calling

For variant calling of *BRCA* mutations, we used the BWA bioinformatic pipeline (v.0.7.15-r1142-dirty) with the -M option for DNA sequence alignment (22). For single nucleotide variants (SNVs), variant calling was performed by two callers: Freebayes (v1.3.6) (23) and Mutect2 (24) from the GATK package (4.1.4.1) (25). Variants were filtered based on allelic frequency (higher than 15%), genomic coverage (above 35) and the number of supporting reads (more than 5). UMI data were processed using the standard FGBio UMI pipeline (v0.8.1) and the following submodules: GroupReadByUMI with “adjacency” as strategy option, CallMolecularConsensus and FilterConsensusRead.

We implemented a proprietary machine learning approach that classifies variants according to the joint recommendations by the American College of Medical Genetics and Genomics (ACMG) and the Association for Molecular Pathology (AMP) (26), further referred to as “ACMG guidelines”). Class 4 and 5 variants were selected for *BRCA* pathogenic variant calling. To determine HRD status, SeqOne Genomics has devised a proprietary solution based on a machine learning approach which includes the number of large genomic alterations, the number of parental copy losses, and amplifications at two gene locations (*CCNE1, RAD51B*) in addition to *BRCA* mutations. This model provides a probability that the tumor sample is HRD-positive: if the probability is greater than 50%, the sample is classified as positive. The data on the HRD and *BRCA* status obtained with Myriad MyChoice were retrieved from the results of the PAOLA-1 cohort (9).

### Statistical Analysis

Progression-free survival curves were generated using the Kaplan-Meier method. An unstratified Cox proportional hazard regression model was used to determine the hazard ratios and associated 95% confidence intervals (95% CI). Confidence intervals for overall percentage agreement and percentage values were calculated with an exact binomial test. Confidence intervals for survival time point estimates were computed using a log transformation (survfit function default from the survival R package). All statistical analyses were performed using the R statistical software, version 4.2.3 (27).

### Data availability

The data can be requested from the clinical and translational departments of the non-profit ARCAGY-GINECO collaborative group.

## Results

### Patients and tumors

The baseline characteristics of the 368 patients included in this study are provided in Table 1). For most of the patients, the primary tumor was located in the ovaries (86% of the patients), with a majority of the tumors being FIGO stage III (73% of all tumors) and of the high-grade serous histological subtype (94%). Nearly three quarters of the patients (72%) had the ECOG Performance Status of 0. These characteristics were similar to those of the entire PAOLA-1 cohort (806 patients).

**Table 1:** Baseline characteristics of the 368 PAOLA-1 patients included in this study.

| Characteristic | All<br>(n=368) | olaparib + bevacizumab arm<br>(n=247) | bevacizumab arm<br>(n=121) |
| --- | --- | --- | --- |
| Patient age | 61 (53 – 67) | 61 (54 – 67) | 61 (52 – 67) |
| ECOG at baseline† |  |  |  |
| 0 | 264 (72%) | 174 (70%) | 90 (74%) |
| 1 | 101 (27%) | 71 (29%) | 30 (25%) |
| Unknown | 3 (1%) | 2 (1%) | 1 (1%) |
| Primary tumor location |  |  |  |
| Ovary | 315 (86%) | 205 (83%) | 110 (91%) |
| Peritoneum | 27 (7%) | 22 (8.9%) | 5 (4.1%) |
| Fallopian tube | 26 (7%) | 20 (8.1%) | 6 (5.0%) |
| FIGO stage |  |  |  |
| III | 270 (73%) | 184 (74%) | 86 (71%) |
| IV | 98 (27%) | 63 (26%) | 35 (29%) |
| Histological subgroup |  |  |  |
| High-grade endometrioid | 11 (3%) | 6 (2.4%) | 5 (4.1%) |
| High-grade serous | 347 (94%) | 238 (96%) | 109 (90%) |
| Other | 10 (3%) | 3 (1.2%) | 7 (5.8%) |
| History of cytoreductive surgery |  |  |  |
| Interval debulking | 114 (31%) | 80 (32%) | 34 (28%) |
| Upfront surgery | 242 (66%) | 161 (65%) | 81 (67%) |
| No surgery | 12 (3%) | 6 (3%) | 6 (5%) |
| Response to first-line chemotherapy¶ |  |  |  |
| Complete Response | 65 (18%) | 44 (18%) | 21 (17%) |
| No evidence of disease | 217 (59%) | 146 (59%) | 71 (59%) |
| Partial Response | 86 (23%) | 57 (23%) | 29 (24%) |
| Normal serum CA-125 level | 330 (90%) | 219 (89%) | 111 (92%) |
| BRCA status by Myriad |  |  |  |
| 0 | 258 (70%) | 173 (70%) | 85 (70%) |
| 1 | 109 (30%) | 74 (30%) | 35 (29%) |
| Unknown | 1 (0%) | 0 | 1 (1%) |
| BRCA status by SeqOne |  |  |  |
| 0 | 257 (70%) | 173 (70%) | 84 (70%) |
| 1 | 107 (29%) | 72 (29%) | 35 (29%) |
| Unknown | 4 (1%) | 2 (1%) | 2 (1%) |
| HRD status by Myriad |  |  |  |
| Negative | 144 (39%) | 101 (41%) | 43 (36%) |
| Positive | 197 (54%) | 128 (52%) | 69 (57%) |
| Unknown | 27 (7%) | 18 (7.3%) | 9 (7.4%) |
| HRD status SeqOne |  |  |  |
| Negative | 164 (45%) | 111 (45%) | 53 (44%) |
| Positive | 200 (54%) | 134 (54%) | 66 (55%) |
| Unknown | 4 (1%) | 2 (1%) | 2 (1%) |
| Median (IQR); n (%) |  |  |  |

†Eastern Cooperative Oncology Group (ECOG) performance status ranges from 0 to 5, with higher values reflecting greater disability.
¶No evidence of disease was defined as no measurable or assessable disease after cytoreductive surgery plus no radio-logic evidence of disease and a normal CA-125 level after chemotherapy. Clinical complete response was defined as the disappearance of all measurable or assessable disease and normalization of CA-125 levels, Partial response was defined as radiologic evidence of disease, an abnormal CA-125 level, or both.

### Comparison between the SeqOne and Myriad HRD testing results

First, we compared the tumor *BRCA* status determined using the automatic variant calling approach implemented in the SeqOne platform to the results obtained with the Myriad MyChoice assay. The results are summarised in Table 2. We found that 70% of tumors were *BRCA*-wildtype (BRCA-wt) using both assays (257 of 368 tumors with the SeqOne approach 258 of 368 with Myriad). Pathogenic *BRCA* variants were found in 29% of tumors (107 of 368 using SeqOne and 30% with Myriad (109 of 368). Only 1% of SeqOne results were inconclusive (4/368) due to failed DNA amplification, while for the Myriad assay, it was below 1% (1/368). The overall concordance of *BRCA* calling results between Myriad and SeqOne was 96.4% (351/364 samples with concordant results; 95% CI: 94%-98%). Among the thirteen samples with discordant results, seven were found to be *BRCA*-mutant with Myriad but *BRCA*-wt with SeqOne, while it is the other way around for the other six samples.

**Table 2:** *BRCA* and HRD status obtained with SeqOne and Myriad MyChoice assay for the tumor samples of the 368 PAOLA-1 patients included in this study.

| Characteristic | ALL (n=368) | olaparib plus beva arm (n=247) | bevacizumab arm (n=121) |
| --- | --- | --- | --- |
| *BRCA* status Myriad |  |  |  |
| 0 | 258 (70%) | 173 (70%) | 85 (70%) |
| 1 | 109 (30%) | 74 (30%) | 35 (29%) |
| Unknown | 1 (0%) | 0 | 1 (1%) |
| *BRCA* status by SeqOne |  |  |  |
| 0 | 257 (70%) | 173 (70%) | 84 (70%) |
| 1 | 107 (29%) | 72 (29%) | 35 (29%) |
| Unknown | 4 (1%) | 2 (1%) | 2 (1%) |
| HRD status by Myriad |  |  |  |
| Negative | 144 (39%) | 101 (41%) | 43 (36%) |
| Positive | 197 (54%) | 128 (52%) | 69 (57%) |
| Unknown | 27 (7%) | 18 (7.3%) | 9 (7.4%) |
| HRD status by SeqOne |  |  |  |
| Negative | 164 (45%) | 111 (45%) | 53 (44%) |
| Positive | 200 (54%) | 134 (54%) | 66 (55%) |
| Unknown | 4 (1%) | 2 (1%) | 2 (1%) |
| n (%) |  |  |  |

Then, we compared the HRD status obtained with the two assays, assuming that HRD-positive samples corresponded to samples with a *BRCA* mutation and/or genomic instability (GIS ≥42 with Myriad or Proba(HRD)≥50% with SeqOne). The SeqOne-determined HRD status relies on sWGS, which was successfully obtained for 364 of 368 tumors, with a mean sequencing coverage of 2.98X (first quartile: 1.13; third quartile: 5.12). We found that 54% of the samples were HRD-positive according to both assays (200 of 368 using the SeqOne platform and 197/368 using the Myriad assay). However, the rates of inconclusive results differed between the two assays, with a higher rate for Myriad: 7% (27 of 368 samples) versus 1% for SeqOne (4/368). All these results are summarized in Table 2. The overall result concordance between the two tests was 95.0% (321 of 338 samples with concordant results; 95% CI: 92%-97%) (Table 3). Assuming that Myriad My Choice provides the reference HRD status, SeqOne assay has a Positive Percentage Agreement (PPA) of 95% (185 out of 195 positive samples; 95% CI; 90%-97%) and a Negative Percentage Agreement (NPA) of 95% also (136 out of 143 negative samples; 95% CI; 89%-98%). Among the 17 samples with discordant results, 10 were found to be HRD-positive with Myriad MyChoice only and 7 only with SeqOne sWGS. Amongst those discordant samples, there was an increased proportion of samples with intermediate values of HRD probabilities by the SeqOne assay; 53% (9/17; 95% CI, 29%-76%) of the discordant samples had intermediate HRD probability values, between 25% and 75%, whereas only 11% (35/321; 95% CI, 8%-15%) of the concordant samples had probability values in that range.

**Table 3:**
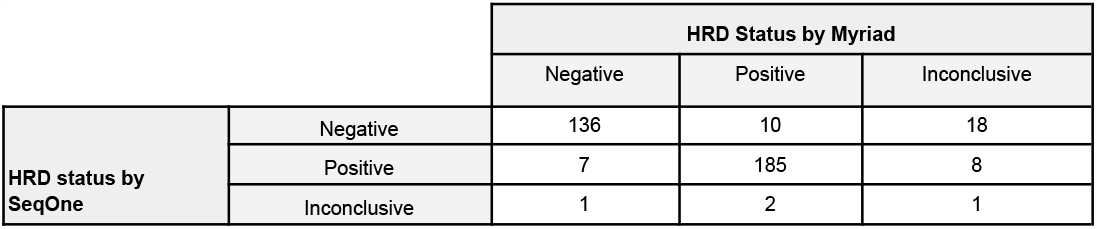
Concordance of homologous recombination deficiency (HRD) tumor status results obtained with the SeqOne HRD and the Myriad MyChoice test for the 368 patients from the PAOLA-1 cohort included in this study.

### SeqOne HRD test results and patient survival

We found a significantly longer PFS for patients with *BRCA*-mutant tumors in the olaparib arm (olaparib + bevacizumab treatment) compared to the control arm (placebo plus bevacizumab), regardless of which test was used to call *BRCA* pathogenic variants (Hazard Ratio (HR): 0.28; 95% CI: 0.16-0.47 with SeqOne; HR: 0.26; 95% CI: 0.16-0.45 with Myriad;Figures 1A and 2). Over a half of these patients (54%) in the olaparib arm did not progress after 60 months of follow-up (95% CI: 44%-68% for both tests). In the control arm, this proportion was 16% (95% CI: 8%-35%) when using SeqOne results of *BRCA* variant calling and 17% (95% CI: 8%-36%) based on the Myriad test.

**Figure 1:**
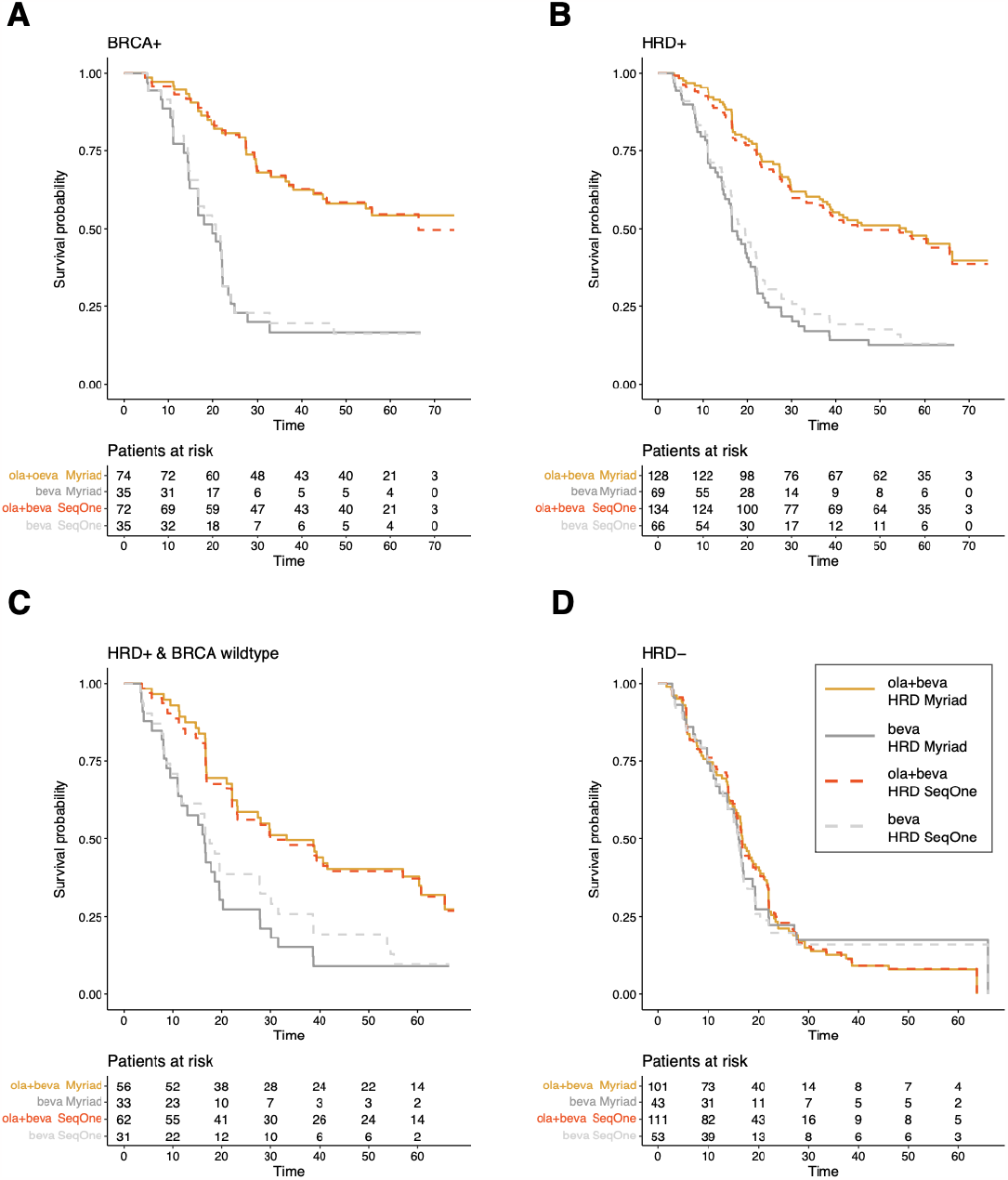
Survival analysis for ovarian cancer patients comparing the olaparib plus bevacizumab arm (Ola+Beva) to the control bevacizumab arm (Beva) for different patient subgroups. Panel A. Patients with tumors carrying a pathogenic *BRCA* variant (*BRCA*+), Panel B. Patients with HRD-positive (HRD+) tumors, Panel C. Patients with HRD-positive and BRCA-wildtype tumors where *BRCA* status has been determined using SeqOne according to the ACMG guidelines, Panel D. Patients with HRD-negative (HRD-) tumors.

**Figure 2:**
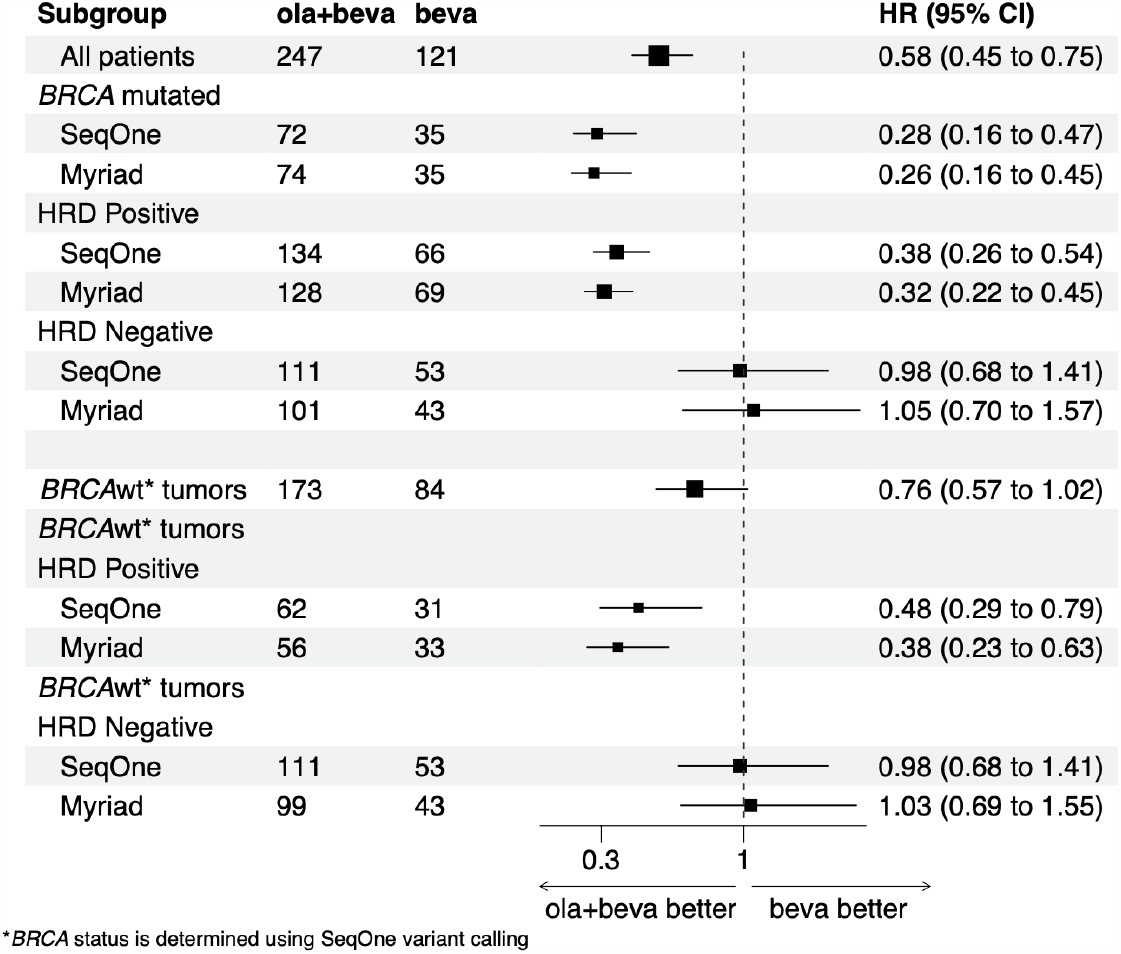
Cox proportional hazard analysis for ovarian cancer patients treated with olaparib and bevacizumab or with bevacizumab only. The patients were stratified by tumor *BRCA* mutation and homologous recombination deficiency (HRD) status, determined with the MyriadMyChoice or the SeqOne assay. Square sizes for the hazard ratios are proportional to the number of patients in each subgroup. The dashed line corresponds to the point of no survival differences between patients in the two arms. Values below 1 indicate a benefit for patients receiving olaparib in addition to bevacizumab. The analysis restricted to the BRCAwt individuals (five estimates of hazard ratio in the bottom part of the plot) has been done by determining *BRCA* status with SeqOne implementation of ACMG guidelines.

Among patients with HRD-positive tumors, PFS was significantly longer in the olaparib arm compared to the control arm, regardless of whether we used SeqOne sWGS test (HR: 0.38; 95% CI: 0.26-0.54) or Myriad MyChoice (HR: 0.32; 95% CI: 0.22-0.45) to define the subgroup (Figures 1B and 2). Stratifying the analysis according to the genomic coverage of the sWGS samples, we found that hazard ratios were significantly below 1 both for samples with a genomic coverage below 1X (HR: 0.25; 95% CI: 0.08-0.71) and for those with the coverage above 1X (HR: 0.40; 95% CI: 0.27-0.59). Among patients with tumors identified as HRD-positive by SeqOne, 47% (95% CI: 39%-56%) did not progress after 60 months of follow-up in the olaparib arm compared to 13% (95%CI: 7%-24%) in the control arm. When we used the Myriad test to determine HRD status, these figures were 48% (95% CI: 40%-58%) and 12% (95% CI: 7%-24%; Figure 1B), respectively.

Among patients with BRCA-wt tumors, those with HRD-positive tumors had significantly longer PFS in the olaparib arm than in the control arm (Figures 1C and 2). The results were in a similar range whether the HRD positivity was determined using the SeqOne sWGS test (HR:0.48; 95% CI: 0.29-0.79) or Myriad MyChoice (HR:0.38; 95% CI: 0.23-0.63). A total of 41% of patients (95%CI: 29%-55%) in the olaparib arm did not progress at 40 months compared to 19% in the control arm (95% CI: 8%-38%) when using the SeqOne test to determine the HRD status, whereas with the Myriad test, these percentages were of 43% (95% CI: 29%-57%) and 9% (95% CI: 2%-25%;Figure 1B). PFS did not significantly differ between the two arms for patients with HRD-negative tumors (HRD-/*BRCAwt*), whether using the SeqOne test, or the Myriad assay to identify the HRD-negative subgroup (HR: 0.98; 95% CI: 0.68-1.41; and HR: 1.05; 95% CI: 0.70-1.57, respectively; Figures 1D and 2). After 60 months of follow-up, the proportion of progression-free patients was 8% (95% CI: 4%-16%) in the olaparib arm, regardless of which test was used to determine the HRD status. In the control arm, it was16% (95% CI, 8%-30%) when using the SeqOne test and 17% (95% CI, 9%-34%) with the Myriad assay.

Next, we analyzed survival for the 26 patients for whom we obtained tumor HRD results with the SeqOne assay, whereas Myriad provided inconclusive results. A total of 18 patients were HRD-according to SeqOne assay and 8 were HRD+. Comparing the olaparib and control arm, we found non-significant differences between arms for both the sWGS HRD-positive subgroup (HR: 0.24; 95% CI: 0.02-2.31) and the sWGS HRD-negative subgroup (HR:1.27; 95% CI: 0.35-4.67), with the point estimates of the hazard ratios that follows and expected direction.

### Reproducibility and repeatability analyses

To validate the utility of the SeqOne test for decentralized in-house use, we conducted reproducibility and repeatability experiments. To test the reproducibility of HRD results, we compared HRD probabilities and statuses obtained for 94 PAOLA-1 samples with two different sequencing systems: NextSeq 550 and NovaSeq 6000. We found the same results of *BRCA* pathogenic variant calling with the two sequencing systems: 30 out of 94 samples carried a *BRCA* pathogenic variant. The HRD results were concordant for 90 of 94 samples. For three of them, the SeqOne HRD probabilities obtained following sequencing with the NovaSeq 6000 system were between 45% and 65% (Supplementary Figure 1). For the fourth sample with discordant results, the HRD probabilities largely differed between the two sequencing systems: 85% with the NextSeq 550 versus 6% with the NovaSeq 6000.

We additionally tested repeatability of *BRCA* variant detection with the SeqOne assay. To this end, the library of 47 PAOLA-1 samples was prepared and sequenced twice in the same laboratory. We found the same *BRCA* status for all samples in both sequencing runs, with the detection of 21 pathogenic *BRCA* variants in each run.

## Discussion

The PAOLA-1 trial showed that adding olaparib to bevacizumab as the maintenance treatment of high-grade ovarian cancer patients with a positive HRD status increased PFS from 17.7 to 37.2 months and the 5-year overall survival (OS) rate from 42.2% to 55.2% when compared to placebo plus bevacizumab (9,28). Centralized testing approach, such as the FDA-approved Myriad MyChoice solution, has been available for routine HRD status analysis. However, not all patients can benefit from a central testing approach, mainly because it is costly. To enable a broader access to olaparib, one must address the challenge of providing affordable and in-house HRD testing that conforms to national guidelines.

The SeqOne sWGS testing approach provides an affordable solution based on a limited 1X genomic coverage yielding results that are largely concordant with those of the Myriad centralized test. We found a 95% overall result concordance between the SeqOne decentralized approach and the Myriad centralized test. The 95% value is in the range of values reported for comparisons between various in-house decentralized testing solutions and the centralized Myriad test, varying from 87% for the AmoyDx HRD assay to 97% for the Myriad decentralized test, with intermediate concordance rates for SOPHiA DDM HRD (90%) and Ilumina TSO500 (95%) (11,18,29,30). Consistent with the high result concordance between the SeqOne solution and the centralized Myriad test, we found that survival outcomes did not differ whether we used Myriad or the SeqOne sWGS test to define HRD subgroups and that approximately half of the patients had HRD-positive tumors, as reported for the entire PAOLA -1 cohort (9).

The SeqOne sWGS test yielded only 1% of inconclusive results – a rate which was lower than obtained with the Myriad centralized test (7%) in this subset of 368 patients from the PAOLA-1 trial. The rate of inconclusive results with the Myriad assay was even higher in the entire PAOLA cohort and reached 18% (9). Independent evidence about the rate of undetermined HRD status results obtained with Myriad MyChoice arised from the PRIMA trial where 15% of inconclusive Myriad HRD results were obtained for newly diagnosed advanced ovarian cancer patients (5).

Inconclusive HRD results are problematic for several reasons, including the importance of HRD results in guiding maintenance therapy decisions in the frontline setting, and the cost of HRD tests. In this study we showed that the SeqOne sWGS HRD test could be clinically useful in patients where the Myriad test gave inconclusive results. Reducing the rate of inconclusive HRD results marks a substantial progress to better guide maintenance therapy in the frontline setting.

By providing an affordable low coverage whole genome solution, the SeqOne sWGS HRD test can contribute to increasing the availability of HRD testing, especially in countries where HRD testing in a central laboratory abroad is not available. Increasing HRD testing rate will allow additional patients to improve their survival.

## Acknowledgments

The authors would like to thank Aude Lasfargues, Déborah Cardoso, and Christine Montoto-Grillot who provided support for the processing and analysis of the PAOLA-1 samples.

## Supplementary material

**Supplementary Figure 1:**
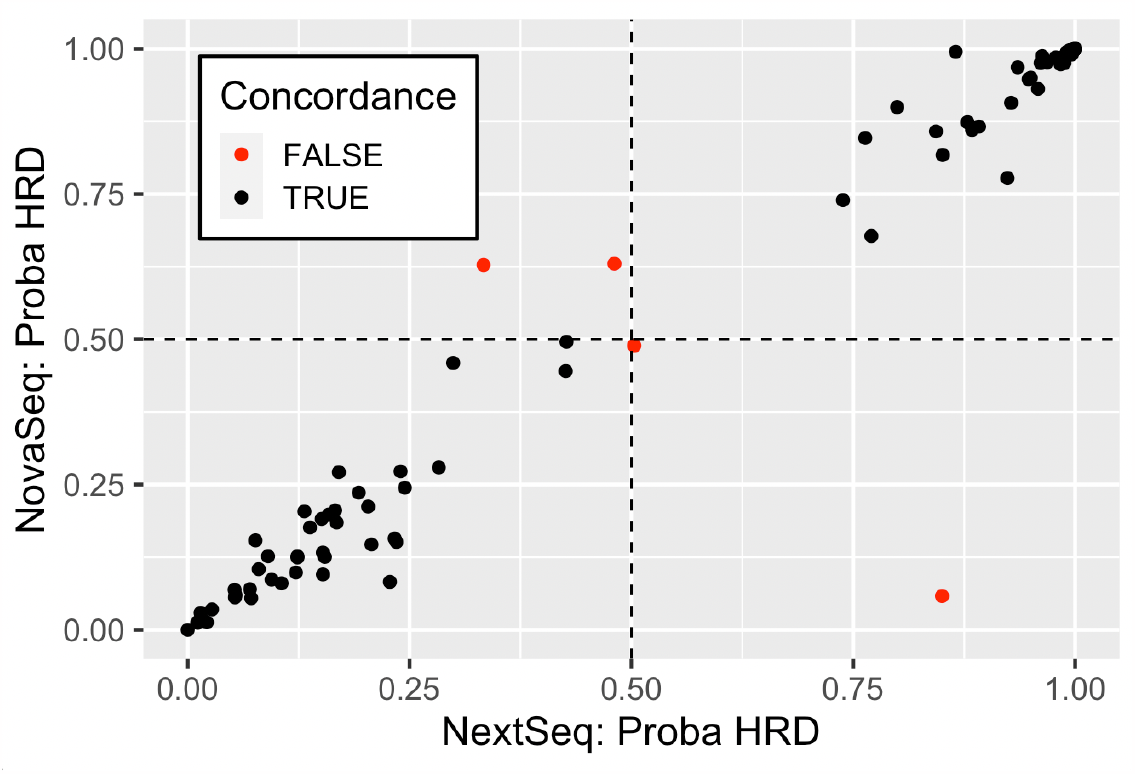
Comparison of the HRD probabilities obtained with the NovaSeq 6000 and NextSeq 550 sequencing systems. A total of 90 out of 94 provided consistent HRD status results.

